# Antifolate resistance markers and complexity of infection in children with sickle cell anaemia receiving malaria chemoprevention in Uganda: a molecular sub-study of the CHEMCHA randomised trial

**DOI:** 10.64898/2026.11.08.26367568

**Authors:** Winnie Nambatya, Moses Ocan, Erick Jacob Okek, Grace Ndeezi, Dennis Kalibbala, Thomas Katairo, Freddie Bwanga, Thandile Nkosi-Gondwe, Joanita Kirikumwino, John Ssenkusu, Stephen Tukwasibwe, Robert Opoka, Feiko O. ter Kuile, Kamija S. Phiri, Richard Idro

## Abstract

**Background:** Sickle cell anaemia (SCA) is a major public health problem in Uganda. Monthly sulfadoxine-pyrimethamine (SP) remains the standard malaria chemoprevention regimen despite widespread antifolate resistance in East Africa. We assessed Pfdhfr and Pfdhps resistance mutations and the complexity of infection (COI) among children with SCA receiving malaria chemoprevention.

**Methods:** This prospective study of antifolate resistance was nested within the CHEMCHA randomised controlled trial comparing monthly SP with weekly dihydroartemisinin-piperaquine (DP) for malaria chemoprevention. Participants were children with SCA aged 6 months to 15 years and attending Jinja and Kitgum hospitals in Uganda. Dried blood spots were collected at enrolment, study closeout, and during febrile episodes. Parasite DNA from microscopy-confirmed clinical malaria samples was genotyped by amplicon sequencing for Pfdhfr (N51I, C59R, S108N, and I164L) and Pfdhps (A437G, K540E, A581G, A613S, and A613T). COI was estimated using MOIRE.

**Results:** Complete Pfdhfr/Pfdhps haplotypes were resolved for 7-28 isolates per site and time point, and COI was estimated for 67 samples. The quintuple mutant (Pfdhfr IRN/Pfdhps GE) accounted for 85.7-100% of isolates across sites and time points; A613S and A613T were not detected. Pfdhfr I164L was detected in 2/9 isolates in Jinja at baseline and 1/8 at endline. The Pfdhps A581G mutant allele was absent at baseline (0/44 isolates) but was found in 1/8 isolates in Jinja and 3/34 in Kitgum at endline, including quintuple-plus-581G (sextuple) haplotypes in 1/8 and 2/27 isolates, respectively. Most infections were monoclonal (51/67, 76.1%).

**Conclusions:** The high prevalence of quintuple antifolate-resistant haplotypes and the detection of higher-level resistance markers, including emerging sextuple mutants carrying Pfdhfr I164L and Pfdhps A581G, call into question the continued use of SP for chemoprevention in children with SCA in Uganda and support evaluation of alternative regimens.

## Background

Sickle cell anaemia (SCA) is a major public health problem, with more than 300,000 affected births annually worldwide, approximately 75% of which occur in sub-Saharan Africa [1]. Global projections suggest that this burden will increase to 400,000 births per year by 2050 [2]. In Uganda, an estimated 15,000–20,000 infants are born with SCA each year [3], and 50%–90% of affected children die before the age of two years [4]. Beyond early mortality, children with SCA experience recurrent complications, including painful vaso-occlusive crises, haemolytic episodes, and severe anaemia, many of which are precipitated by febrile illnesses such as malaria [5].

Although sickle cell trait (HbAS) confers partial protection against *Plasmodium falciparum* malaria [6], children with SCA remain at substantially increased risk of death from malaria [5]. Malaria contributes to morbidity in this population by exacerbating chronic anaemia, a determinant of poor clinical outcomes, and precipitating severe SCA complications. Malaria chemoprevention is therefore a critical component of comprehensive SCA management and has the potential to reduce severe malaria-related morbidity, hospitalisation, blood transfusion requirements, and mortality.

In Uganda, malaria chemoprevention with monthly SP is the standard of care in children with SCA [1]. However, prolonged SP use across East Africa has been associated with the emergence and selection of drug-resistant *P. falciparum* [2]. Over 90% of *P. falciparum* isolates in the East African region carry mutations in the dihydrofolate reductase (*Pfdhfr*) and dihydropteroate synthetase (*Pfdhps*) genes associated with reduced SP efficacy [3–6]. The quintuple mutant haplotype comprising *Pfdhfr* mutations N51I, C59R, and S108N together with *Pfdhps* mutations A437G and K540E is an established molecular marker of SP treatment failure [7, 8]. Some preventive benefit may remain in settings where the prevalence of the Pfdhps K540E mutation is below 50% [9]. Yet, the continued recommendation of SP for chemoprevention in Ugandan children with SCA, despite well-documented regional resistance, leaves an important gap between policy and the available evidence.

Despite widespread use, few data characterise molecular markers of antimalarial resistance in children with SCA. It remains unclear whether resistance profiles in this population are comparable to those observed in the general paediatric population, or whether chemoprevention in this group may selectively drive distinct resistance patterns. These evidence gaps have direct implications for malaria control policy in this high-risk group. Unlike the general paediatric population, in whom SP exposure is intermittent, children with SCA in Uganda receive monthly SP continuously for years. Sustained drug pressure may itself select for resistant parasites, and any loss of efficacy carries a disproportionate clinical cost in a group already at high risk of severe malaria. This study therefore determined the molecular markers of antimalarial resistance in children with SCA receiving malaria chemoprevention in Uganda, to inform malaria prevention strategies and policy in this population.

## Methods

### Study design

This was a nested prospective study of antifolate resistance in children with SCA in Uganda. The study was nested within a randomized, double-blind, parallel-group, placebo-controlled clinical trial of weekly DP versus monthly SP for malaria chemoprevention in children with SCA (CHEMCHA trial) [10].

### Study setting and target population

The parent trial was conducted at two hospitals in Uganda (Jinja Regional Referral Hospital [RRH] and Kitgum General Hospital) and two hospitals in Malawi from April 2021 to May 2023[10, 11]. This substudy was restricted to the two Ugandan sites, both of which lie in areas with high-grade P. falciparum antifolate resistance.

### Sample size

The sample size for the parent trial was calculated to provide 90% power (two-sided α=0.05) to detect a 50% reduction in the incidence of clinical malaria, from an assumed 0.2 episodes per person-year in the SP arm to 0.1 episodes per person-year in the DP arm [11]. This required 824 person-years of follow-up in approximately 548 participants [10]. Because this was a secondary analysis of malaria parasites collected within the trial, no separate sample size calculation was performed; instead, all malaria-positive samples on microscopy were included.

### Sampling procedure and selection of respondents

Participants in the main study [10] were 725 children aged 6 months to 15 years with a diagnosis of SCA on haemoglobin electrophoresis or isoelectric focusing. Of these, the 438 children enrolled at the two Ugandan sites were eligible for this sub-study. Children with other known chronic diseases, those receiving daily cotrimoxazole prophylaxis, or those participating in another trial were excluded.

### Data collection process

The trial procedures have been described previously [10]. Briefly, at enrolment, between 17 April 2021 and 30 May 2022, sociodemographic information and medical history were recorded, and a physical examination was performed [11]. Participants were randomly assigned in a 1:1 ratio to one of two malaria chemoprevention regimens using a computer-generated block randomisation sequence with randomly varying block sizes of 4, 6, or 8, stratified by site and weight category [10]. To maintain blinding, participants in each arm received one active regimen alongside a matched placebo for the alternative regimen: thus, those assigned to the DP arm received weekly active DP and monthly placebo SP, while those assigned to the SP arm received monthly active SP and weekly placebo DP [11]. Participants who were unwell at enrolment were initially resuscitated and enrolled once clinically stable [11].

Participants attended scheduled clinics with their caregivers every 2 months to receive their next doses of study medication, and unscheduled visits if they were unwell. At each study visit, participants receiving SP were provided with two additional doses to cover the subsequent two months, while participants receiving DP were provided with doses sufficient to cover 8 weeks; these additional doses were dispensed on the last day of the preceding scheduled visit. Participants also attended the clinic for all sick visits, including episodes of febrile illnesses. The study provided standard-of-care clinical services for all unscheduled visits. Parents or guardians were instructed on home management of children with SCA and on bringing children to the study clinic after identifying signs requiring medical attention [10, 11]. Participants who missed scheduled follow-up visits were contacted by phone and visited at home if required [10]. Participants were admitted to hospital if they developed severe complications of SCA.

### Sample collection and processing

Samples confirmed for malaria by microscopic examination of blood smears were included in the study. For every positive microscopy finding, a sample was stored on filter paper (Whatman 3MM) for molecular analyses [11]. The blood-blotted filter paper was left to dry at room temperature before storage at 4 °C, awaiting molecular analysis. This procedure was repeated for all recruited participants. Dried blood Spots (DBS) were packaged according to the principles of triple packaging and transported to the National Malaria Reference and Research Laboratory (NMRRL) using the National Health Laboratory Diagnostics System (NHLDS) hub transportation system [12]. The primary container was the DBS cards on which blood was spotted. DBS was wrapped in zip-lock bags (secondary containers) alongside desiccants and sterile cotton gauze. Together with the filled Laboratory investigation forms and ice packs, the zip-lock bag was put inside a cool box (tertiary container). The entire package was handed over to the NHLDS hub transport system.

#### Genotyping drug resistance genes

Parasite genomic DNA from all microscopy-positive samples was extracted from blood spots using Tween-20/Chele× 100 as previously described [13]. Briefly, 6-mm diameter discs were punched from dried blood spots into 1.5-mL microcentrifuge tubes and incubated in 0.05% PBS-T [Tween 20 (Sigma Aldrich P9416) in 1× PBS (Corning 21-040-CV)] overnight at 4 °C. Samples then underwent centrifugation at 13,000 rpm for 1 min; the PBS-T was aspirated and replaced with 1 mL of 1× PBS, and the sample was incubated at 4 °C for 30 min. Following 5 min of centrifugation at 13,000 rpm and 4 °C, the supernatant was aspirated, and 150 µL of 7% Chele× 100 (Bio-Rad 1422822) in water was added. The sample was then incubated with shaking at 95 °C for 10 min. Following centrifugation at 13,000 rpm for 10 min, the supernatant containing extracted DNA was transferred to 1.5-mL sample storage tubes, avoiding transfer of any remaining Chelex beads. Extracted DNA was stored at −20 °C.

Parasite density was estimated using quantitative PCR targeting the *var*ATS gene, as previously described [13]. Samples positive by *var*ATS were selected for genotyping. Library preparation was performed using the *Plasmodium falciparum* Public Health Sequencing Tool (PfPHAST) panel, the smaller panel within the Multiplexed Amplicons for Drugs, Diagnostics, Diversity, and Differentiation using High-Throughput Targeted Resequencing (MAD4HatTeR) panel [14], which comprises two primer pools, M1.1 (45 targets) and M2.1 (15 targets), including 15 drug resistance genes with loci in *crt*, *mdr1*, *dhfr*, *dhps*, and *kelch13*. Samples, including positive and negative controls, were pooled and sequenced on the Illumina MiSeq platform using MiSeq v2 flow cells. Samples with <50% of amplicons having >100 reads were re-prepped, and those with >50% but <75% of amplicons having >100 reads were repooled and resequenced to increase coverage, as previously described [14].

### Data management and analysis

Data were collected and recorded at the point of contact using a combination of paper-based and electronic record forms [11], using the Research Electronic Data Capture (REDCap™) system, locally hosted. Data quality was ensured through comprehensive training of the study team, standardized questionnaire administration and data collection procedures, and good clinical data management and laboratory practices. Built-in range, logic, and consistency checks were incorporated into the electronic system to minimize data entry errors.

Bioinformatic analyses were performed using the ampSeq pipeline, as previously described [18], with downstream analyses conducted in R version 4.2.2. Genotypes were called when an allele had ≥50 reads and a within-sample allele frequency (WSAF) ≥1%. Samples carrying both wild-type and mutant alleles were classified as mixed, and mutant alleles with a WSAF below [n]% were confirmed using a repeat library preparation. Complexity of infection (COI) was estimated using MOIRE, based on the median posterior COI metric, as previously described [20], and rounded to the nearest integer for presentation. Statistical analyses were performed using Stata version 18. Baseline demographic and clinical characteristics were summarised using means and standard deviations (SDs) for continuous variables and frequencies and percentages for categorical variables, with summaries stratified by study site. The primary outcome was the presence of polymorphisms in the *Pfdhfr* and *Pfdhps* genes. Samples collected at enrolment were classified as baseline samples, and those collected at study closeout as endline samples. Genotype prevalence was expressed as the percentage of successfully genotyped samples by site and time point.

### Ethics approval and consent to participate

Written informed consent was obtained from a parent or caregiver, and assent from children aged 8 years or older. The parent trial was approved by the Makerere University School of Medicine Research and Ethics Committee (2020-103), Uganda National Council for Science and Technology (HS709ES), the Liverpool School of Tropical Medicine Research Ethics Committee (19.105), the Regional Committee for Medical and Health Research Ethics, Western Norway (30992), and the National Health Sciences Research Committee in Malawi (19/11/2442).

## Results

### Baseline characteristics of study participants

In total, 438 children with SCA were enrolled at the two Ugandan sites, 248 at Jinja RRH and 190 at Kitgum Hospital, of whom 222 were randomly assigned to DP and 216 to SP. The mean age was 7.3 (SD 3.9) years, and just over half (52.3%, 229/438) of the children were male (Table 1).

**Table 1.**
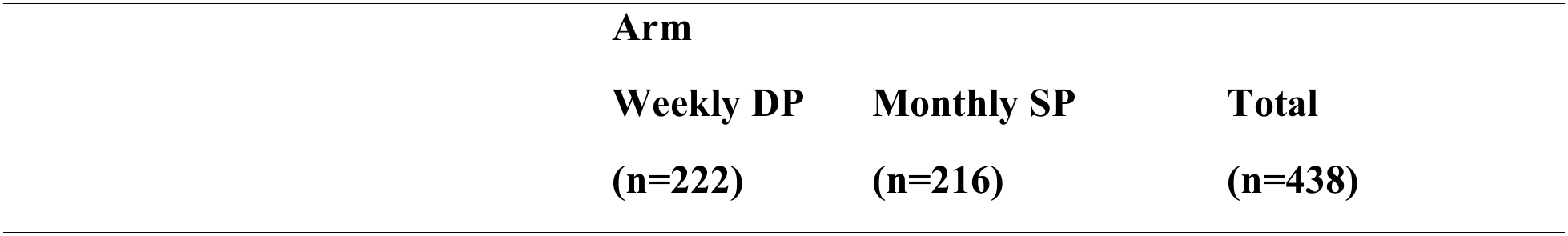
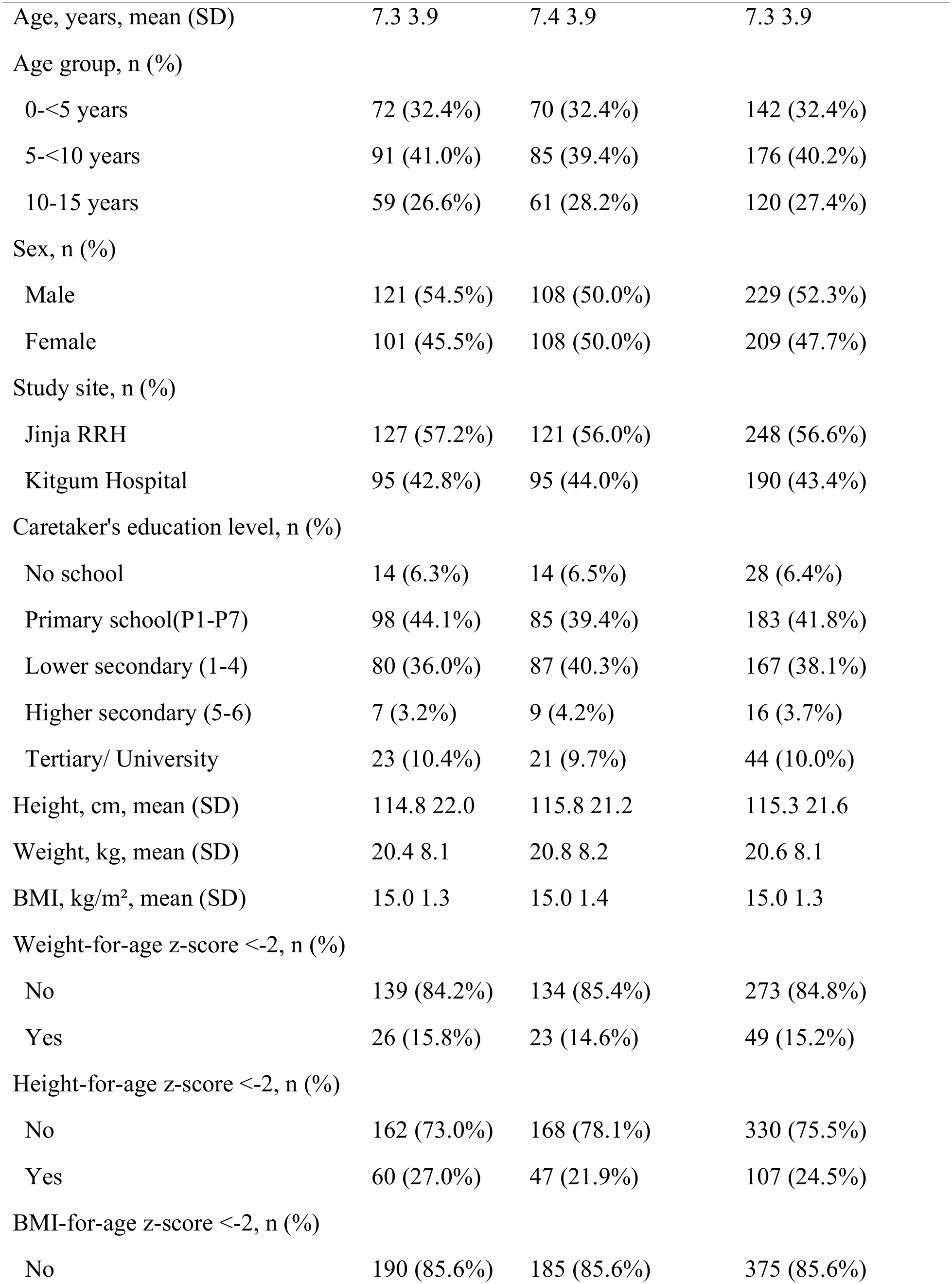
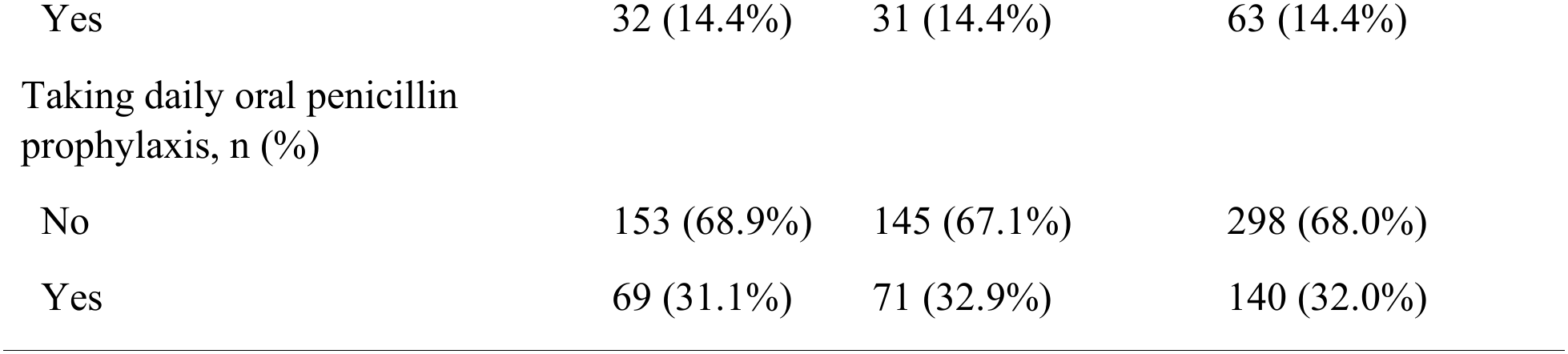
Baseline characteristics of study participants.

|  | **Arm** |  |  |
|---|---|---|---|
|  | **Weekly DP**<br>**(n=222)** | **Monthly SP**<br>**(n=216)** | **Total**<br>**(n=438)** |
| Age, years, mean (SD) | 7.3 3.9 | 7.4 3.9 | 7.3 3.9 |
| Age group, n (%) |  |  |  |
| 0-<5 years | 72 (32.4%) | 70 (32.4%) | 142 (32.4%) |
| 5-<10 years | 91 (41.0%) | 85 (39.4%) | 176 (40.2%) |
| 10-15 years | 59 (26.6%) | 61 (28.2%) | 120 (27.4%) |
| Sex, n (%) |  |  |  |
| Male | 121 (54.5%) | 108 (50.0%) | 229 (52.3%) |
| Female | 101 (45.5%) | 108 (50.0%) | 209 (47.7%) |
| Study site, n (%) |  |  |  |
| Jinja RRH | 127 (57.2%) | 121 (56.0%) | 248 (56.6%) |
| Kitgum Hospital | 95 (42.8%) | 95 (44.0%) | 190 (43.4%) |
| Caretaker's education level, n (%) |  |  |  |
| No school | 14 (6.3%) | 14 (6.5%) | 28 (6.4%) |
| Primary school(P1-P7) | 98 (44.1%) | 85 (39.4%) | 183 (41.8%) |
| Lower secondary (1-4) | 80 (36.0%) | 87 (40.3%) | 167 (38.1%) |
| Higher secondary (5-6) | 7 (3.2%) | 9 (4.2%) | 16 (3.7%) |
| Tertiary/ University | 23 (10.4%) | 21 (9.7%) | 44 (10.0%) |
| Height, cm, mean (SD) | 114.8 22.0 | 115.8 21.2 | 115.3 21.6 |
| Weight, kg, mean (SD) | 20.4 8.1 | 20.8 8.2 | 20.6 8.1 |
| BMI, kg/m², mean (SD) | 15.0 1.3 | 15.0 1.4 | 15.0 1.3 |
| Weight-for-age z-score <-2, n (%) |  |  |  |
| No | 139 (84.2%) | 134 (85.4%) | 273 (84.8%) |
| Yes | 26 (15.8%) | 23 (14.6%) | 49 (15.2%) |
| Height-for-age z-score <-2, n (%) |  |  |  |
| No | 162 (73.0%) | 168 (78.1%) | 330 (75.5%) |
| Yes | 60 (27.0%) | 47 (21.9%) | 107 (24.5%) |
| BMI-for-age z-score <-2, n (%) |  |  |  |
| No | 190 (85.6%) | 185 (85.6%) | 375 (85.6%) |
| Yes | 32 (14.4%) | 31 (14.4%) | 63 (14.4%) |
| Taking daily oral penicillin prophylaxis, n (%) |  |  |  |
| No | 153 (68.9%) | 145 (67.1%) | 298 (68.0%) |
| Yes | 69 (31.1%) | 71 (32.9%) | 140 (32.0%) |

At enrolment, 15.2% (49/322) of children were underweight (weight-for-age z-score <-2), and 24.5% (107/437) had stunting (height-for-age z-score <-2). Most children, 95.4%, had received at least one dose of pneumococcal vaccination (43.2% complete, 52.2% incomplete). In addition, at enrolment, 78% (341/437) of participants reported always taking SP chemoprevention and a further 58 (13.3%) reported sometimes taking it (Table 1). During follow-up, mean adherence to study medication was 98.8%.[10].

### Effect of baseline parasitaemia on time to clinical malaria and the incidence of clinical malaria among children with SCA receiving malaria chemoprevention during follow-up

The median follow-up time was 16.7 months (IQR: 13.7-18.6 months). Overall, the incidence rate of clinical malaria was 39.2 per 100 person-years (95% CI 34.2-44.6). In the DP arm, the incidence of clinical malaria was 13.0 per 100 person-years (95% CI 9.2-17.9) and 65.6 per 100 person-years (95% CI 56.6-75.7) in the SP arm. These clinical outcomes have been reported for the full trial population elsewhere [10] and are presented here only to describe the population from which the genotyped samples were drawn.

Children with parasites on microscopy at enrolment developed clinical malaria earlier than those without (Fig. 1; log-rank p < 0.001). The cumulative probability of a first episode in the baseline-parasite group (n = 58) increased to approximately 50% by month 5, 64% by month 10, and 93% by month 18. In the group without baseline parasites (n = 379), the cumulative probability increased slowly, to approximately 10% by month 8, 14% by month 10 and 25% by month 19. The number of children still at risk fell sharply over follow-up: 300 of 379 (no baseline parasites) and 21 of 58 (baseline parasites) remained at risk at month 10, and 22 and 1 at month 20. Fig 1.

**Fig 1.**
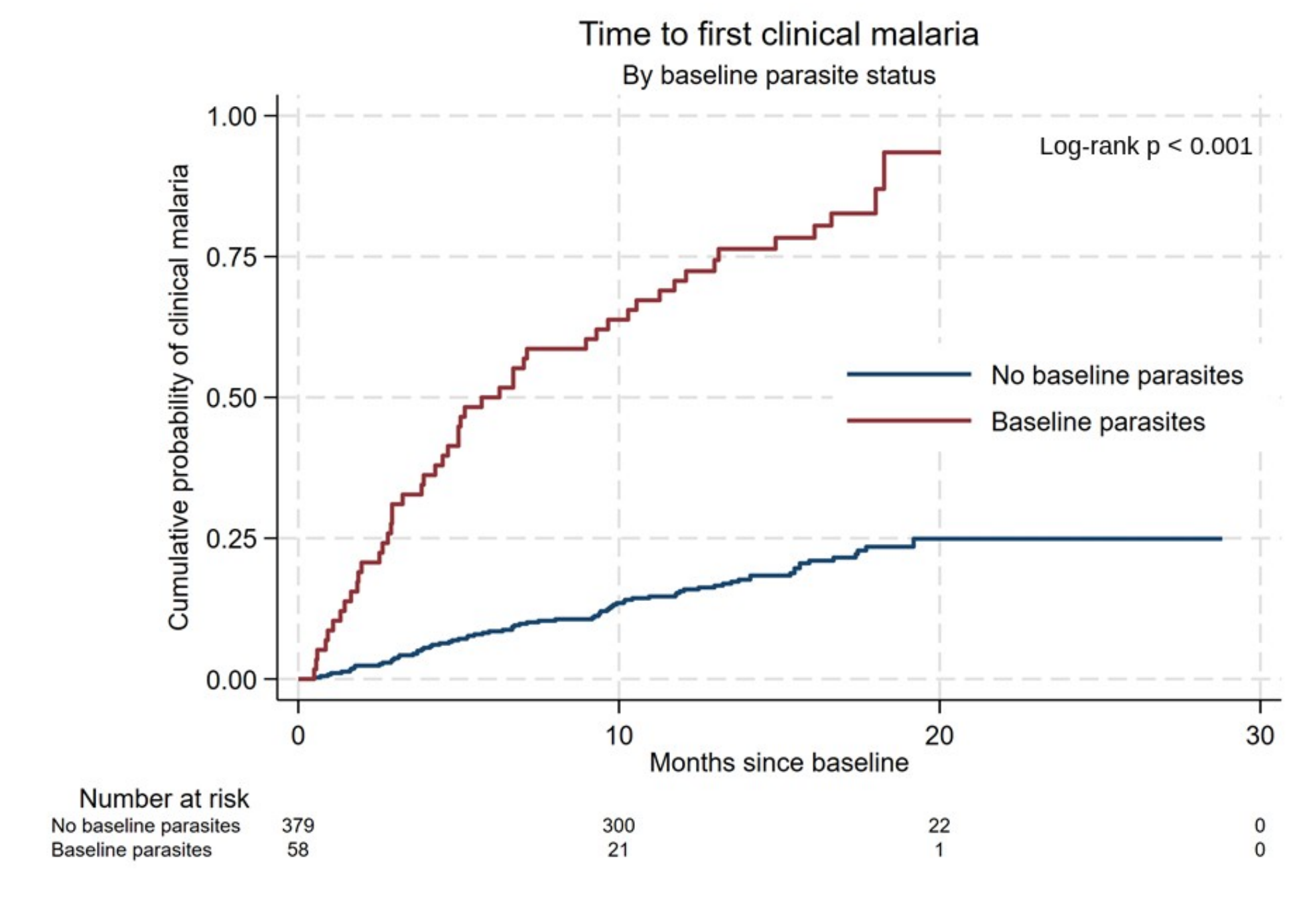
Time to first clinical malaria by baseline parasite status.

### Distribution of parasite density among qPCR-positive participants by study site and study time point

Twelve participants tested positive for malaria parasites on qPCR in Jinja and 9 tested positive at the end of the study. Most had low parasitemia. In Kitgum, a higher proportion, 36/190, tested positive at the start of the study and an even higher proportion (54/190) at study end. Parasite densities were also higher (Figure 2).

**Fig 2.**
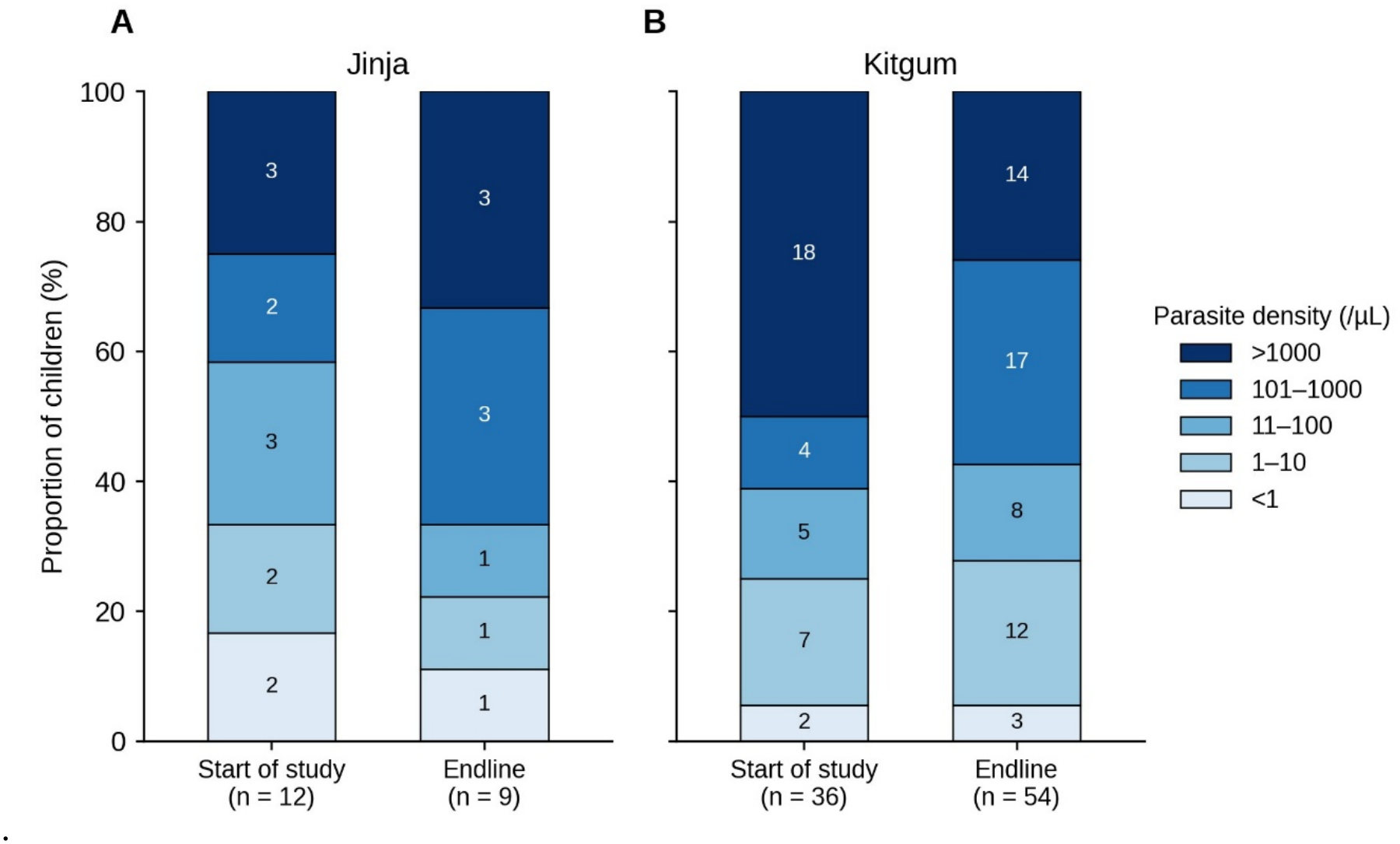
Distribution of parasite density among qPCR-positive participants by study site and study time point. Parasite density results were available for 48 children at the start of the study (12 in Jinja and 36 in Kitgum) and 63 at endline (9 in Jinja and 54 in Kitgum).

### Prevalence of molecular markers of resistance in the Pfdhfr and Pfdhps genes among children with SCA with clinical malaria receiving malaria chemoprevention

The distribution of Pfdhfr and Pfdhps genotypes at the start of the study and at endline is shown in Fig. 3, and the counts for each SNP are given in S1 Table. For Pfdhfr, the N51I and S108N mutants were present in all isolates and at both sites and time points (N51I: 11/11 and 8/8 in Jinja, and 31/31 and 32/32 in Kitgum, at the start of the study and at endline, respectively; S108N: 9/9 and 8/8 in Jinja, and 30/30 and 31/31 in Kitgum). C59R was mutant in all isolates at the start of the study (11/11 in Jinja and 31/31 in Kitgum) and in all isolates in Jinja at endline (8/8); in Kitgum at endline it was mutant in 29/32 isolates, and 3/32 were wild type. The I164L mutation was detected in 2/9 isolates in Jinja at the start of the study and in 1/8 at endline. In Kitgum, it was not detected at the start of the study (0/30), and one isolate at endline had a mixed genotype (1/31).

**Fig. 3.**
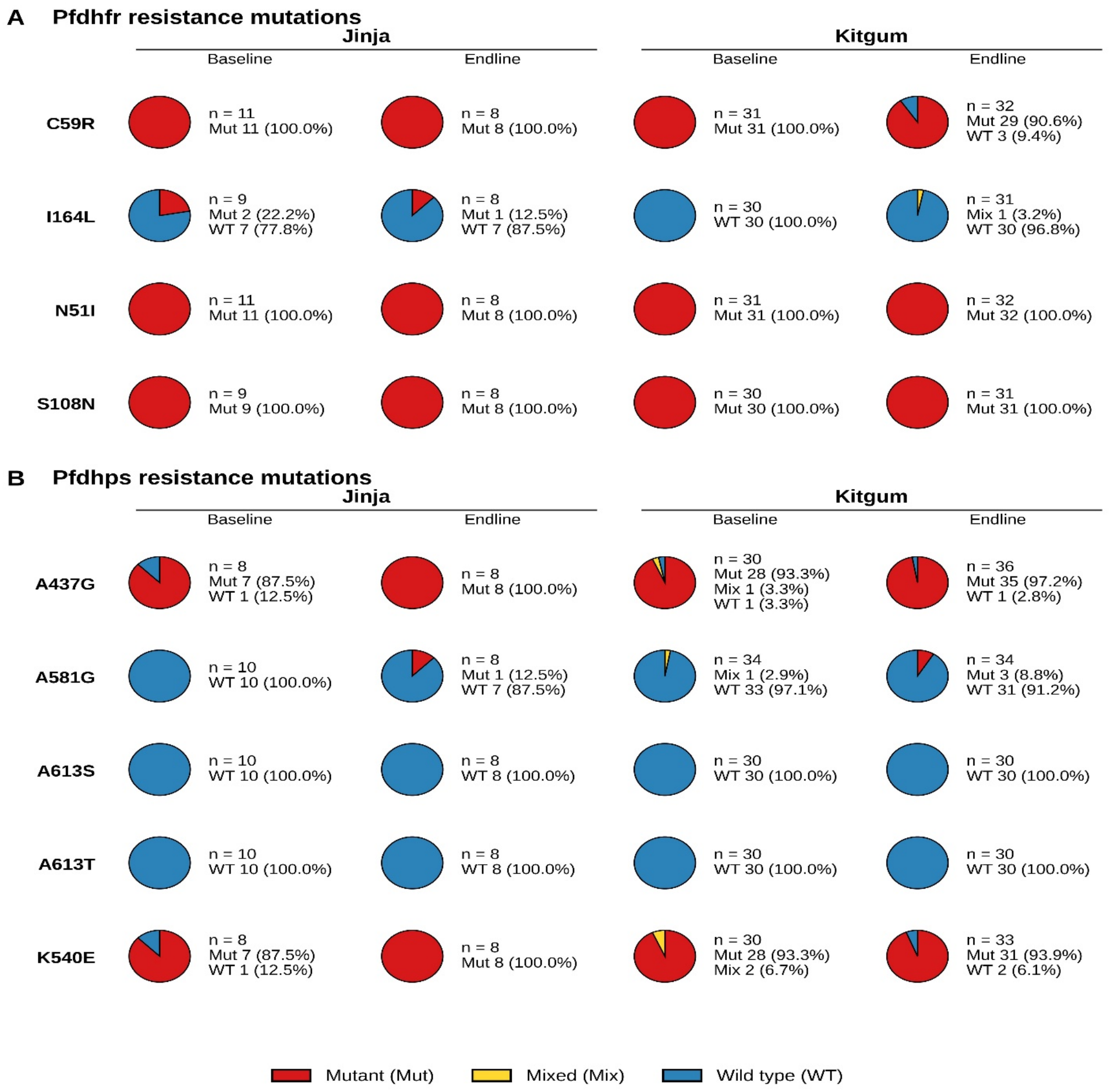
Prevalence of Plasmodium falciparum Pfdhfr and Pfdhps drug-resistance markers at enrolment and at the end of the study in Jinja and Kitgum districts, Uganda.

For Pfdhps, the K540E mutation was detected in 7/8 isolates in Jinja and 28/30 in Kitgum at the start of the study (with two mixed genotypes in Kitgum), and in 8/8 and 31/33 isolates at endline, respectively. The A437G mutation was detected in 1/8 isolates in Jinja and 1/30 in Kitgum at the start of the study (with one further mixed genotype in Kitgum), and in 0/8 and 1/36 isolates at endline, respectively. The A581G mutation was not detected at the start of the study although one isolate in Kitgum had a mixed genotype (1/34). At endline, the A581G mutant allele was detected in 1/8 isolates in Jinja and 3/34 isolates in Kitgum. A613S and A613T were wild type in all isolates at both sites and time points (Jinja: 10/10 at the start of the study and 8/8 at endline; Kitgum: 30/30 at both time points).

### Frequencies of Plasmodium falciparum dhfr/dhps haplotypes at baseline and endline, stratified by site

The quintuple mutant haplotype (IRN plus GE) predominated at both sites and at both time points (Fig. 4A). In Jinja, it was detected in 6/7 isolates at the start of the study and in 8/8 at endline, and in Kitgum in 25/26 and 24/28, respectively. Other haplotypes were uncommon. They were IRN plus AK in Jinja at the start of the study (1/7,), and ICN plus GE (2/28,), IRN plus AK (1/28), and IRN plus GK (1/28) in Kitgum at endline.

The quintuple plus 581G haplotype, which corresponds to the sextuple mutant, was not detected in Jinja at the start of the study (0/7) but was detected in 1/8 isolates at endline. In Kitgum, it was detected in 1/26 isolates at the start of the study and in 2/27 at endline (Fig. 4B). The quintuple plus 164L haplotype was also detected in 1/7 isolates in Jinja at the start of the study, 1/8 in Jinja at endline, 0/26 in Kitgum at the start of the study, and 1/27 in Kitgum at endline.

**Fig. 4.**
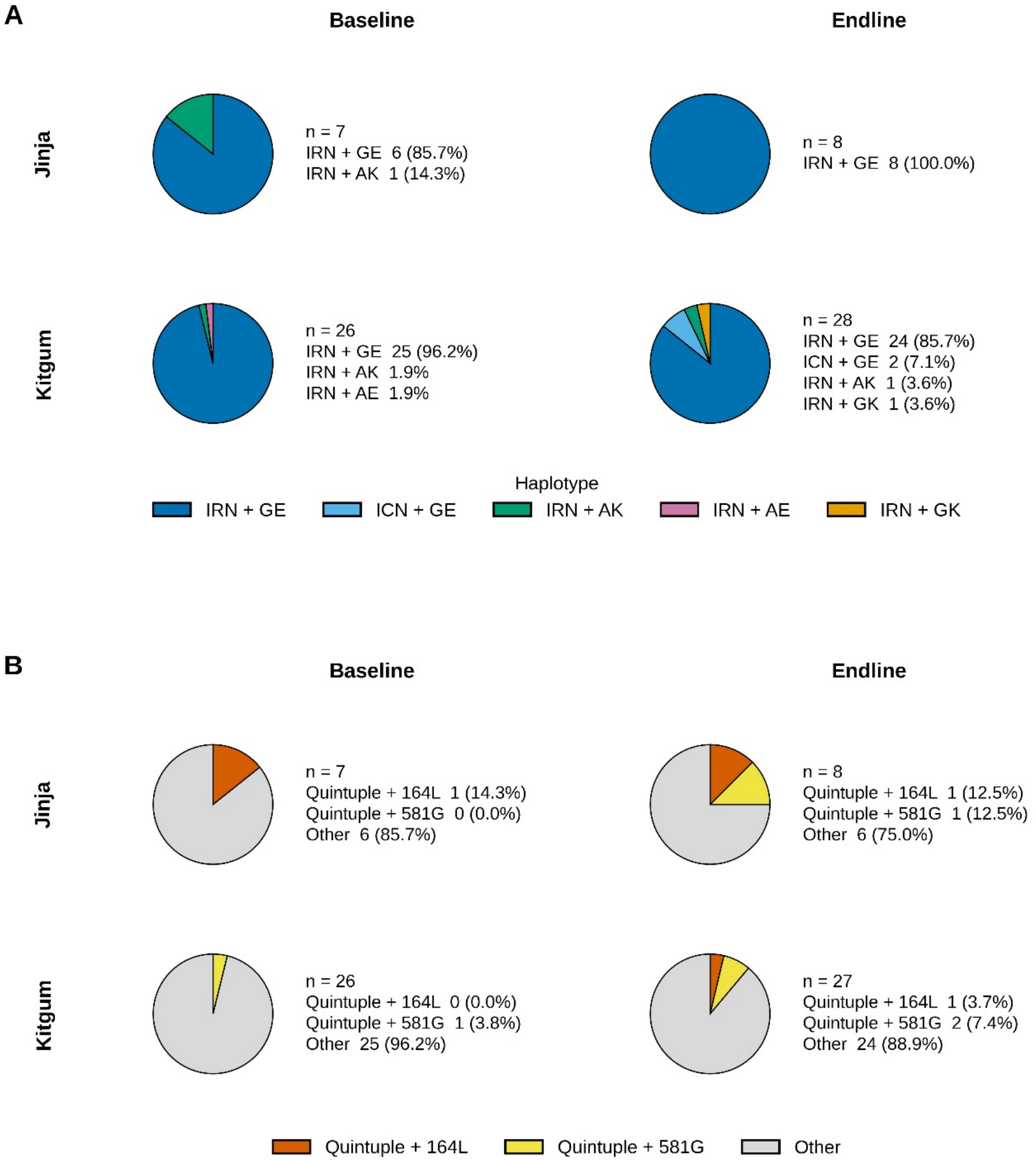
Frequencies of Plasmodium falciparum dhfr/dhps haplotypes at baseline and endline, stratified by site. Haplotype codes combine the Pfdhfr codon 51/59/108 genotype with the Pfdhps codon 437/540 genotype. IRN = Ile51 + Arg59 + Asn108-(N51I/C59R/S108N); ICN = Ile51 + Cys59 (wild-type) + Asn108-(N51I/S108N); GE = Gly437 + Glu540-(A437G/K540E); AE = Ala437 (wild-type) + Glu540-(K540E only); AK = Ala437 (wild-type) + Lys540 (wild-type), wild-type at both codons; GK = Gly437 + Lys540 (wild-type)-(A437G only).

### Distribution of the complexity of infection

Complexity of infection (COI) was determined for 67 samples. Most infections were monoclonal (51/67, 76.1%), followed by infections with a COI of 2 (12/67, 17.9%). COI values of 3 and 4 were each observed in 2/67 samples (3.0%) (Fig. 3).

**Fig. 3a.**
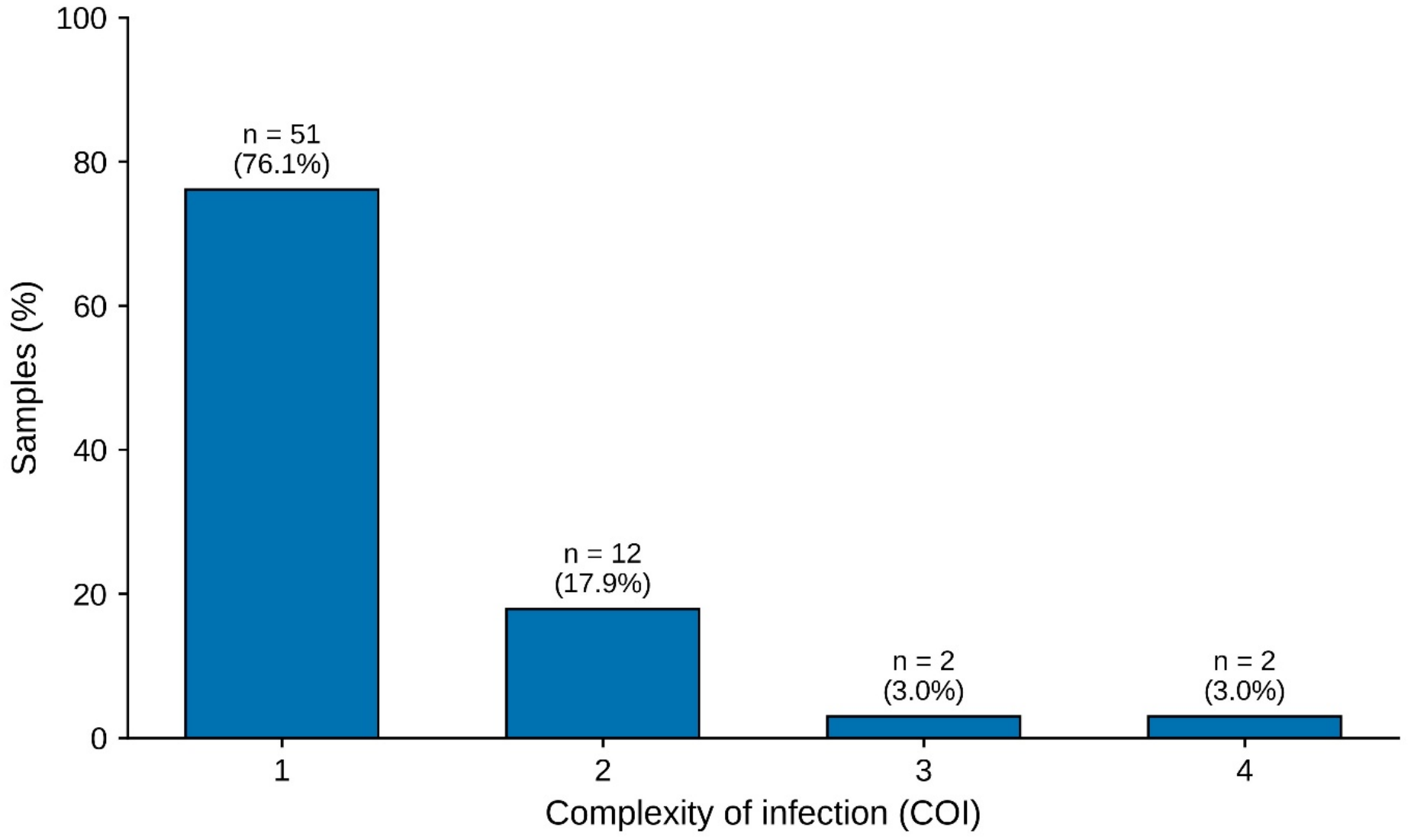
Distribution of the complexity of infection (COI), estimated as the effective multiplicity of infection using MOIRE and presented as integer values, among all successfully genotyped samples from both sites and time points combined

## Discussion

This study characterised molecular markers of antifolate resistance and the COI among *Plasmodium falciparum* parasites infecting children with SCA receiving malaria chemoprevention in Jinja and Kitgum, Uganda, at enrolment and at the end of follow-up. The Pfdhfr N51I, C59R, S108N, and Pfdhps K540E and A437G mutations were present in nearly all successfully genotyped isolates. At the haplotype level, the Pfdhfr IRN/Pfdhps GE quintuple-mutant background predominated at both sites and time points. Additional markers associated with higher-level antifolate resistance, Pfdhfr I164L and Pfdhps A581G, were also detected. These findings are particularly relevant in the context of the parent CHEMCHA trial, in which the incidence of clinical malaria was approximately five times higher among children receiving monthly SP than among those receiving weekly DP [10]. The appearance of A581G during follow-up, in parasites causing breakthrough malaria in children taking monthly SP, is consistent with selection of the most resistant parasites under continuous drug pressure. These data argue for revision of the Ugandan recommendation of monthly SP for children with sickle cell anaemia: weekly DP is the evidence-based alternative in children younger than 5 years receiving penicillin prophylaxis [10]; further evaluation is needed in older children, and continuing SP in this group risks selecting for parasites carrying still higher-level antifolate resistance.

At baseline, a higher proportion of the samples from Kitgum had parasite densities exceeding 1000 parasites/µL, consistent with the higher transmission intensity of that region [15]. At endline, parasite densities in Kitgum shifted toward lower strata, which may indicate partial suppression of high-density infections by chemoprevention. However, the persistence of infections in the 1-10 and 101-1000 parasites/µL strata also suggests ongoing low-grade transmission despite prophylaxis. In Jinja, parasite density distributions were broadly stable at the beginning of the study and endline, consistent with its peri-urban setting.

The prevalence and pattern of *Pfdhfr* and *Pfdhps* mutations observed in this study, including the near-universal quintuple mutant haplotype, are similar to those reported in general paediatric and antenatal populations elsewhere in East Africa [5]. This suggests that children with SCA are not exposed to a distinct resistance profile compared with the wider population, although their continuous use of SP chemoprevention may increase the clinical consequences of existing resistance.

The high prevalence of the *Pfdhfr* N51I, C59R, and S108N mutations, which together constitute the triple mutant (IRN) haplotype conferring high-level pyrimethamine resistance, at both study sites is also consistent with previous evidence showing that these variants are already highly prevalent across East Africa [2, 3, 5]. Their presence at or close to 100% at both time points and at both sites suggests that these mutations are now fixed, or nearly fixed, in the local parasite population. Similarly, the high prevalence of the *Pfdhps* A437G (88-100%) and K540E (88-95%) mutations indicates that the quintuple mutant haplotype is dominant in this population.

Haplotype-level analysis confirmed this pattern. The full quintuple mutant (Pfdhfr IRN/Pfdhps GE) accounted for 86-100% of isolates in Jinja and 86-96% of isolates in Kitgum at baseline and endline, with only a small proportion of isolates carrying partial mutant backgrounds. The presence of the quintuple mutant among children with SCA is particularly concerning because these children receive monthly SP chemoprevention continuously over several years, unlike the general paediatric population, in whom SP exposure is typically intermittent. Sustained drug pressure in this population may reduce the protective benefit of SP and favour the persistence or further selection of resistant parasite populations.

The presence of both I164L and A581G in this population deserves attention because these mutations have been uncommon in Uganda [6] and mediate high-level SP resistance [16]. In a 2024 survey of children with malaria in northern, north-eastern and eastern Uganda, I164L was found in a few samples, and A581G was reported as a mixed genotype [17]. By contrast, the Pfdhps A581G mutant allele, which was not detected at baseline, was found at endline at both sites. One possible explanation is selection by continued SP exposure, since SP given for intermittent preventive treatment in pregnancy in Uganda has been associated with selection of I164L and A581G [5, 18]. The presence of the quintuple and sextuple haplotype is particularly relevant in children with SCA, who receive monthly SP chemoprevention continuously over several years, in contrast to the more intermittent SP exposure typically experienced by the general paediatric population. Given that a *Pfdhps* K540E prevalence above 50% has been associated with reduced preventive efficacy of SP [9], the levels observed here (88-95%) suggest that SP chemoprevention may already provide suboptimal protection for children with SCA in Uganda, and there is a need to revise the malaria chemoprevention policies.

The predominance of monoclonal infections across both sites may reflect suppression of polyclonal parasite populations by continuous chemoprevention. Previous studies in Uganda have reported polyclonal infections in more than 50% of samples across sites of varying transmission intensity, in community-based cohorts of children (6 months-10 years) [19]. The lower COI observed in this chemoprevention cohort is therefore consistent with the capacity of antimalarial prophylaxis to reduce parasite diversity and select for persistence of single dominant clones [20].

Taken together, these findings suggest that most circulating parasites in this population carry a molecular profile associated with SP treatment failure. This raises important concerns about the continued use of SP as the standard chemoprevention regimen for children with SCA in Uganda and supports re-evaluation of the current malaria policy.

### Limitations

This study has several limitations. It was a secondary analysis nested within a clinical trial, and the sample sizes at individual sites and time points were relatively small, which may have limited the precision of the haplotype-frequency estimates. The analysis was also restricted to the Ugandan trial sites; therefore, the findings may not be generalisable to settings with different transmission intensities, resistance profiles, or chemoprevention histories. In addition, the data are not linked to individual clinical outcomes; therefore, no inferences can be drawn about the association between a specific haplotype and the risk of clinical malaria.

## Conclusions

Molecular markers of SP resistance are near universal among children with SCA receiving malaria chemoprevention in Uganda. The quintuple mutant haplotype predominated at both sites, Pfdhfr I164L was present from baseline, and Pfdhps A581G, undetected at baseline, appeared at endline in parasites causing breakthrough malaria, consistent with selection of sextuple mutants under continuous monthly SP. The profile is no different from that of the wider parasite population, but its consequences are, because these children take SP every month for years and are at high risk of severe malaria. Taken with the five-fold higher incidence of clinical malaria in the SP arm of the parent trial, these findings indicate that monthly SP is no longer adequate chemoprevention in this group. Weekly DP should replace it in children younger than 5 years receiving penicillin prophylaxis; its efficacy and safety should be established in older children, and molecular surveillance should continue under whichever regimen is adopted.

## Acknowledgements

We thank study participants for their involvement in the CHEMCHA study. We also thank the CHEMCHA trial team for patient clinical management and data collection. In addition, we acknowledge the support from Professor Bjarne Robberstad and the National Malaria Reference and Research Laboratory (NMRRL), Kampala, for their contributions to molecular analysis.

## Availability of data and materials

The data underlying the results presented in this study are held on the servers of Global Health Uganda and will be made available upon institutional approval from the administration at Global Health Uganda (Plot 667 Mawanda Road, Kampala, Uganda; telephone +256 393 516707). Data access is controlled as a way of maintaining participant confidentiality in line with the Ethical Review Board requirements.

## Supporting information

S1 Table. Prevalence of Pfdhfr and Pfdhps antifolate resistance-associated mutations by study site and time point, Jinja and Kitgum districts, Uganda.

## References

1. Ministry of Health Uganda. Uganda Clinical Guidelines 2023. Available from: https://library.health.go.ug/sites/default/files/resources/Uganda%20Clinical%20Guidelines%202023.pdf.

2. Asua V, Conrad MD, Aydemir O, Duvalsaint M, Legac J, Duarte E, et al. Changing prevalence of potential mediators of aminoquinoline, antifolate, and artemisinin resistance across Uganda. The Journal of Infectious Diseases. 2021;223(6):985–94.

3. Mbonye AK, Birungi J, Yanow SK, Shokoples S, Malamba S, Alifrangis M, et al. Prevalence of Plasmodium falciparum resistance markers to sulfadoxine-pyrimethamine among pregnant women receiving intermittent preventive treatment for malaria in Uganda. Antimicrobial Agents and Chemotherapy. 2015;59(9):5475–82. doi: 10.1128/aac.00507-15.

4. Gikunju SW, Agola EL, Ondondo RO, Kinyua J, Kimani F, LaBeaud AD, et al. Prevalence of pfdhfr and pfdhps mutations in Plasmodium falciparum associated with drug resistance among pregnant women receiving IPTp-SP at Msambweni County Referral Hospital, Kwale County, Kenya. Malar J. 2020;19(1):190. Epub 20200524. doi: 10.1186/s12936-020-03263-z. PubMed PMID: 32448228; PubMed Central PMCID: PMCPMC7247205.

5. Kizza J, Katairo T, Kakuru A, Nsengimaana B, Esilu T, Wiringilimaana I, et al. Recurrent Plasmodium falciparum Parasitemia and Drug Resistance Mutations During Intermittent Preventive Treatment of Malaria in Pregnancy in Uganda. The Journal of Infectious Diseases. 2026. doi: 10.1093/infdis/jiag074.

6. Katairo T, Asua V, Semakuba FD, Nsengimaana B, Kiyaga S, Hathaway N, et al. Sustained high prevalence of multiple antimalarial drug resistance markers in Uganda in 2023– 2024. Antimicrobial Agents and Chemotherapy. 2026:e00361–26.

7. Abdullah NR, Norahmad NA, Jelip J, Sulaiman LH, Mohd Sidek H, Ismail Z, et al. High prevalence of mutation in the Plasmodium falciparum dhfr and dhps genes in field isolates from Sabah, Northern Borneo. Malar J. 2013;12:198. Epub 20130612. doi: 10.1186/1475-2875-12-198. PubMed PMID: 23758930; PubMed Central PMCID: PMCPMC3706343.

8. Tchuenkam PVK, Ngum LN, Ali IM, Chedjou JPK, Nji AM, Netongo PM, et al. Plasmodium falciparum dhps and dhfr markers of resistance to sulfadoxine-pyrimethamine five years (2016-2020) after the implementation of seasonal malaria chemoprevention in Cameroon. Wellcome Open Res. 2024;9:323. Epub 20240620. doi: 10.12688/wellcomeopenres.22347.1. PubMed PMID: 39649624; PubMed Central PMCID: PMCPMC11624434.

9. Organization WH. Intermittent preventive treatment for infants using sulfadoxine-pyrimethamine (SP-IPTi) for malaria control in Africa: IMPLEMENTATION FIELD GUIDE. World Health Organization, 2011.

10. Idro R, Nkosi-Gondwe T, Opoka R, Ssenkusu JM, Dennis K, Tsirizani L, et al. Weekly dihydroartemisinin–piperaquine versus monthly sulfadoxine–pyrimethamine for malaria chemoprevention in children with sickle cell anaemia in Uganda and Malawi (CHEMCHA): a randomised, double-blind, placebo-controlled trial. The Lancet Infectious Diseases. 2025;25(6):643–55. doi: 10.1016/S1473-3099(24)00737-0.

11. Nkosi-Gondwe T, Robberstad B, Opoka R, Kalibbala D, Rujumba J, Galileya LT, et al. Dihydroartemisinin-piperaquine or sulphadoxine-pyrimethamine for the chemoprevention of malaria in children with sickle cell anaemia in eastern and southern Africa (CHEMCHA): a protocol for a multi-centre, two-arm, double-blind, randomised, placebo-controlled superiority trial. Trials. 2023;24(1):257. doi: 10.1186/s13063-023-07274-4.

12. Kiyaga C, Sendagire H, Joseph E, McConnell I, Grosz J, Narayan V, et al. Uganda’s new national laboratory sample transport system: a successful model for improving access to diagnostic services for early infant HIV diagnosis and other programs. PloS one. 2013;8(11):e78609.

13. Katairo T, Asua V, Nsengimaana B, Tukwasibwe S, Semakuba FD, Wiringilimaana I, et al. Performance of molecular inversion probe DR23K and Paragon MAD4HatTeR Amplicon sequencing panels for detection of Plasmodium falciparum mutations associated with antimalarial drug resistance. Malaria journal. 2025;24(1):188.

14. Aranda-Díaz A, Neubauer Vickers E, Murie K, Palmer B, Hathaway N, Gerlovina I, et al. Sensitive and modular amplicon sequencing of Plasmodium falciparum diversity and resistance for research and public health. Scientific Reports. 2025;15(1):10737.

15. Uganda National Malaria Control Division UBoS, ICF,. Uganda Malaria Indicator Survey 2018–19. 2020.

16. Kublin JG, Dzinjalamala FK, Kamwendo DD, Malkin EM, Cortese JF, Martino LM, et al. Molecular markers for failure of sulfadoxine-pyrimethamine and chlorproguanil-dapsone treatment of Plasmodium falciparum malaria. The Journal of infectious diseases. 2002;185(3):380–8.

17. Olupot-Olupot P, Paasi G, Katairo T, Alunyo JP, Nakiyemba A, Ocen GG, et al. Comprehensive analysis of molecular markers linked to antimalarial drug resistance in Plasmodium falciparum in Northern, Northeastern and Eastern Uganda. Malaria journal. 2025;24(1):190.

18. Nayebare P, Asua V, Conrad MD, Kajubi R, Kakuru A, Nankabirwa JI, et al. Associations between malaria-preventive regimens and Plasmodium falciparum drug resistance-mediating polymorphisms in Ugandan pregnant women. Antimicrobial agents and chemotherapy. 2020;64(12):10.1128/aac.01047-20.

19. Mwesigwa A, Ocan M, Cummings B, Musinguzi B, Kiyaga S, Kiwuwa SM, et al. Plasmodium falciparum genetic diversity and multiplicity of infection among asymptomatic and symptomatic malaria-infected individuals in Uganda. Tropical Medicine and Health. 2024;52(1):86.

20. Liljander A, Chandramohan D, Kweku M, Olsson D, Montgomery SM, Greenwood B, et al. Influences of intermittent preventive treatment and persistent multiclonal Plasmodium falciparum infections on clinical malaria risk. PLOS ONE. 2010;5(10):e13649.

